# Titin-Truncating variants Predispose to Dilated Cardiomyopathy in Diverse Populations

**DOI:** 10.1101/2024.01.17.24301405

**Authors:** John DePaolo, Marc Bornstein, Renae Judy, Sarah Abramowitz, Shefali S. Verma, Michael G. Levin, Zoltan Arany, Scott M. Damrauer

## Abstract

**Importance:** The effect of high percentage spliced in (hiPSI) *TTN* truncating variants (TTNtvs) on risk of dilated cardiomyopathy (DCM) has historically been studied among population subgroups defined by genetic similarity to European reference populations. This has raised questions about the effect of TTNtvs in diverse populations, especially among individuals genetically similar to African reference populations.

**Objective:** To determine the effect of TTNtvs on risk of DCM in diverse population as measured by genetic distance (GD) in principal component (PC) space.

**Design:** Cohort study

**Setting:** Penn Medicine Biobank (PMBB) is a large, diverse biobank.

**Participants:** Participants were recruited from across the Penn Medicine healthcare system and volunteered to have their electronic health records linked to biospecimen data including DNA which has undergone whole exome sequencing.

**Main Outcomes and Measures:** Risk of DCM among individuals carrying a hiPSI TTNtv.

**Results:** Carrying a hiPSI TTNtv was associated with DCM among PMBB participants across a range of GD deciles from the 1000G European centroid; the effect estimates ranged from odds ratio (OR) = 3.29 (95% confidence interval [CI] 1.26 to 8.56) to OR = 9.39 (95% CI 3.82 to 23.13). When individuals were assigned to population subgroups based on genetic similarity to the 1000G reference populations, hiPSI TTNtvs conferred significant risk of DCM among those genetically similar to the 1000G European reference population (OR = 7.55, 95% CI 4.99 to 11.42, *P*<0.001) and individuals genetically similar to the 1000G African reference population (OR 3.50, 95% CI 1.48 to 8.24, *P*=0.004).

**Conclusions and Relevance:** TTNtvs are associated with increased risk of DCM among a diverse cohort. There is no significant difference in effect of TTNtvs on DCM risk across deciles of GD from the 1000G European centroid, suggesting genetic background should not be considered when screening individuals for titin-related DCM.

**Key Points:** 

**Question:** Do high percentage spliced in (hiPSI) titin truncating variants (TTNtvs) confer similar levels of risk for dilated cardiomyopathy (DCM) across diverse populations?

**Findings:** In a cohort study that comprised 43,731 individuals of diverse genetic background with electronic health records linked to whole exome sequencing data, hiPSI TTNtvs conferred increased risk of DCM across all individuals irrespective of genetic background as measured by genetic distance from the 1000 Genomes Project European centroid.

**Meaning:** The findings of this study suggest that TTNtvs increase risk of DCM among individuals independent of genetic background and that genetic similarity to a reference population should not play a role in screening for genetic causes of dilated cardiomyopathy.

## Introduction

Titin, the protein encoded by the *TTN* gene, is the largest protein in the human body. It is found in the sarcomere where it spans from the Z-disk to the M-band, and is critical for sarcomere assembly, contraction and relaxation in striated cardiac muscle.^1^ In populations largely composed of individuals genetically similar to the 1000 Genomes Project (1000G) European reference population (EUR), heterozygous *TTN* truncating variants (TTNtvs) that encode for shortened forms of the titin protein have been identified as a common genetic cause of dilated cardiomyopathy (DCM). These TTNtv have been associated with 25% of familial cases of DCM, and 10-20% of sporadic cases.^2–7^ Only variants located in exons that are highly likely to be spliced into adult cardiac *TTN* transcripts, known as high percentage spliced in (hiPSI) variants, are pathogenic.^3^ hiPSI TTNtvs may cause DCM by reducing abundance of full-length TTN protein (haploinsufficiency) and/or through dominant negative effects.^8,9^

We previously demonstrated the association of hiPSI TTNtvs with DCM exclusively in EUR individuals, but were unable to detect an association between hiPSI TTNtvs and DCM among Penn Medicine Biobank (PMBB) participants genetically similar to the 1000G African reference population (AFR; odds ratio [OR] 1.8, 95% CI 0.2 to 13.7, *P*=0.57), or among AFR participants of the Jackson Heart Study.^7^ Recently, a cross-sectional analysis of individuals with DCM demonstrated a statistically significant but attenuated effect of predicted loss-of-function (pLOF) variants in *TTN* among AFR individuals compared to EUR individuals, potentially underscoring the limitations of applying genetic understanding of disease derived from a single ancestry group to diverse populations.^10^

Common and rare genetic risk factors for disease have historically been analyzed among different populations separated by ancestry.^11–14^ This reflects the belief that the artificial grouping of individuals into genetically common cohorts allows improved assignment of genetic risk. However, greater understanding of genetic similarities between groups of individuals combined with the results of genetic admixture within different populations suggests that instead of strict dichotomization into groups, genetic ancestry is better thought of as a continuum of relative similarity.^12^

Here we describe the assessment of DCM risk conferred by hiPSI TTNtvs in a large, diverse biobank. Instead of using genetic similarity to group individuals together, each participant’s genetic distance (GD) from the 1000G European centroid was calculated and risk of DCM was assessed across a range of genetic distances. As a sensitivity analysis, participants were assigned an “population group” according to their genetic similarity to one of the 1000G continental-level reference populations and risk of DCM was stratified by “genetic similar” group. Due to previous research identifying TTNtvs risk factors for atrial fibrillation (Afib),^15–17^ we performed a similar investigation into the link between TTNtvs and Afib.

## Methods

### Study Population

The Penn Medicine BioBank is a genomic and precision medicine cohort comprising participants who receive care in the Penn Medicine health system and who consent to linkage of electronic health records with biospecimens, including 43,731 with DNA which has undergone whole exome sequencing (<u>pmbb.med.upenn.edu</u>). As previously described,^7^ DCM was defined either as ≥2 outpatient or ≥1 inpatient encounters with: 1) the *International Classification of Diseases, 10^th^ Revision* (ICD10) diagnosis code of I42.0; or 2) ICD10 codes I42.8 or I42.9 or the *International Classification of Diseases, Ninth Revision* (ICD9) codes 425.4, 425.8, or 425.9, and mention of “dilated cardiomyopathy” or “DCM” in free text encounter notes. Ischemic cardiomyopathy (ICM) was defined as ≥2 outpatient or ≥1 inpatient encounters with ICD10 codes I24 or I25, or ICD9 codes 411 or 414. Afib was defined as ≥2 outpatient or ≥1 inpatient encounters with ICD10 codes I48, I48.1, I48.2, or I48.9, or ICD9 codes 427.2 or 427.21. A total of 9,020 individuals in PMBB had transthoracic echocardiography (TTE) data available that included left ventricular ejection fraction (LVEF).

### Genetic Data

Whole exome sequencing was performed as previously described by Regeneron Genomics Center.^18^ Individual patient DNA samples were processed and sequenced on the Illumina NovaSeq 6000 (Albany, NY, USA). WeCall variant caller (v2.0.0) was employed for sequence alignment (GRCh38), variant identification, and genotype assignment. Quality control exclusions included sex errors, high rates of heterozygosity (D-statistic > 0.4), low sequence coverage, and genetically identified sample duplicates.

Single nucleotide variants (SNVs) were filtered for a read depth ≥7 and were retained if they either had at least one heterozygous variant genotype with an allele balance ratio ≥0.15, or a homozygous variant genotype.^19^ Insertion-deletion variants (INDELs) were filtered for a read depth ≥10 and either a heterozygous variant genotype with an allele balance ≥0.20, or a homozygous variant genotype.

### TTN Truncating Variant Calls

TTNtvs were filtered for minor allele frequency < 0.001 and selected based on predicted loss of function, truncating variants using ANNOVAR.^20^ Splice site variants were screened for those affecting canonical donor or acceptor splice sites (two bases flanking either exon). Variants were considered hiPSI if percent spliced in was greater than 90% (PSI>0.9).^3^

### Genetic Distance

To consider differences in genetic ancestry along a continuous scale, genetic distance (GD) from the 1000G European centroid was calculated as previously decribed.^14^ Briefly, the 43,731 individuals in the PMBB were projected onto the 1000G^21^ principal component (PC) space. The GD was calculated by taking the Euclidean distance for each individual from the 1000G European centroid using the equation 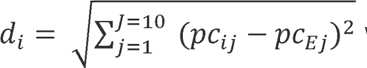 where *pc_ij_* is the *j*th PC of individual *I*, *pc_Ej_* is the 1000G European mean of the *j*th PC, and *J* is set to 10. GD was used to bin each individual in PMBB into deciles from the closest 10% to the European centroid to the furthest 10% from the European centroid.

### Statistical Analysis

Logistic regression was employed to evaluate the association of hiPSI TTNtvs with prevalent DCM adjusting for age and sex. Linear regression was used to assess the effect of hiPSI TTNtvs on minimum left ventricular ejection fraction (LVEF) adjusting for age and sex. Statistical analyses were performed using R (version 4.2.0).

## Results

### Study Population

There were 43,371 participants in the analytic cohort (**Table 1**). The median age at analysis was 57 years (IQR: 45-69 years) and 21,907 (50%) were female. One percent (436 individuals) carried a hiPSI TTNtv, 1,112 individuals (2.5%) had a DCM diagnosis, and 4,920 individuals (11%) had an Afib diagnosis. There were 365 unique TTNtvs identified including 174 stop-gains, 44 frameshifting insertion-deletions, and 147 essential splice site variants.

**Table 1:** Clinical characteristics of individuals in the Penn Medicine Biobank with and without high percentage spliced-in *TTN* truncating variants.

| **Demographic** | **hiPSI TTNtv**<br>**(436)** | **non-hiPSI TTNtv**<br>**(43,295)** | **P-value**<br>**Difference** |
| --- | --- | --- | --- |
| Age, median (IQR) | 56 (45-67) | 57 (45-69) | 0.65 |
| Female Sex, N (%) | 184 (42%) | 21,723 (50%) | <0.001 |
| "Genetically Similar" Group, N (%) |  |  |  |
| EUR | 330 (75%) | 29,626 (68%) | 0.002 |
| AFR | 88 (20%) | 11,048 (26%) | 0.008 |
| AMR | 2 (0.4%) | 564 (1.3%) | 0.12 |
| EAS | 4 (0.9%) | 466 (1.1%) | 0.74 |
| SAS | 5 (1.1%) | 554 (1.3%) | 0.79 |
| Unknown | 7 (1.6%) | 1,037 (2.4%) | 0.28 |
| Dilated Cardiomyopathy, N (%) | 52 (12%) | 1,060 (2.4%) | <0.001 |
| Ischemic Cardiomyopathy, N (%) | 64 (15%) | 5,305 (12%) | 0.13 |
| Atrial Fibrillation, N (%) | 89 (20%) | 4,831 (11%) | <0.001 |

### Calculating genetic distance

GD as the calculated Euclidean distance provides a method to assess the risk of DCM conferred by TTNtvs across a genetically diverse population without relying on artificial grouping based on genetic similarity to a reference panel. Euclidean distance is the distance between two points in n^th^-dimensional space. The vast majority of understanding of the genetic risk of DCM, including that conferred by TTNtvs, was derived from investigation of cohorts primarily comprised of individuals genetically similar to the 1000G European reference population. Integrating the genetic distance from the 1000G European centroid into the analysis of DCM risk among TTNtv carriers accounts for population differences between individuals and the cohort from which genetic understanding of disease has been derived. Therefore, GD provides two useful tools in the current analysis: 1) a method to treat genetic similarity as a continuum rather than an artificially dichotomized value; and 2) a way to test if decreasing genetic similarity indicates a change in TTNtv-derived DCM risk that might support previous findings of differing effects among genetically dissimilar groups.

GD was calculated for each individual in PMBB and genetic diversity by GD was compared to categorical assignment to population subgroups similar to the 1000G reference populations based on PC analysis (**Figure 1A-D**). To demonstrate the extent of genetic admixture and the arbitrary nature of grouping individuals genetically similar to reference populations, PMBB individuals were compared to 1000G reference panel individuals based on PC analysis (**Figure 1E-G**). Individuals were then binned by GD decile; each decile included a range of individuals genetically similar to a 1000G reference population (**Supplemental Table 1**) and PC analysis by GD decile redemonstrated the extent of genetic admixture (**Supplemental Figure 1**). Together these analyses characterize the genetic heterogeneity in the PMBB population.

**Figure 1:**
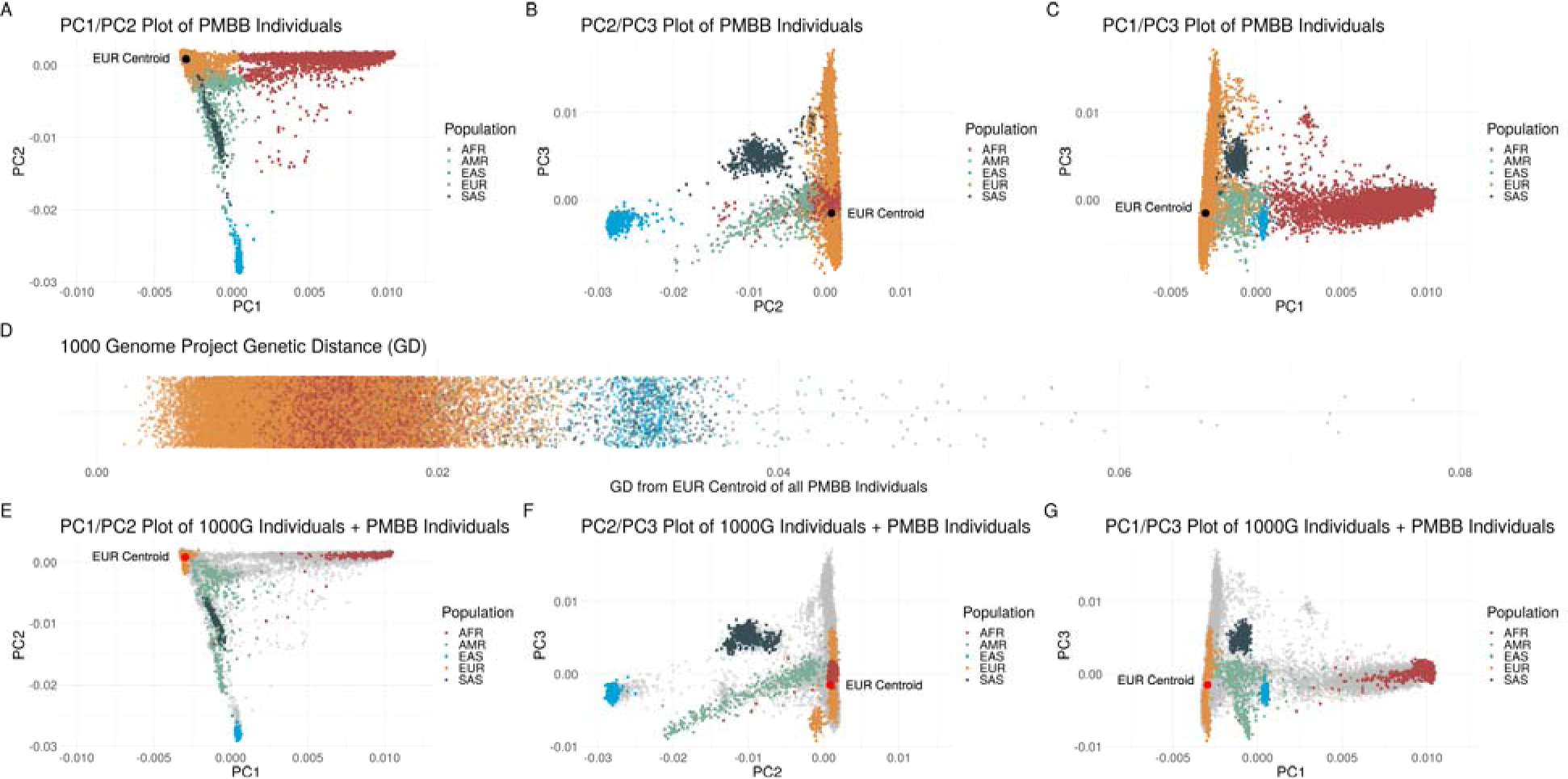
Principal component based clusters of individuals in the Penn Medicine Biobank compared to genetic distance from the 1000 Genomes Project European centroid and compared to continental-level clusters of 1000 Genomes Project individuals demonstrating the degree of overlap of different genetically similar groups. A-C: Discrete labelling of the position of each individual within the Penn Medicine Biobank (PMBB) colored by genetically similar group based on 1000 Genomes Project (1000G) continental-level reference populations in a plot of (**A**) principal component (PC) 1 versus PC2, (**B**) PC2 versus PC3, and (**C**) PC1 versus PC3 with 1000G European centroid (black dot) identified in each plot. (**D**) the genetic distance from the 1000G European centroid of each individual within PMBB again colored by genetically similar group based on 1000 Genomes Project (1000G) continental-level reference populations. **E-G:** Discrete labelling of the position of each 1000G reference panel individuals colored by continental region with PMBB individuals represented by grey dots in a plot of (**E**) PC1 versus PC2, (**F**) PC2 versus PC3, and (**G**) PC1 versus PC3 with 1000G European centroid (red dot) identified in each plot.

### Effect of TTNtvs on risk of DCM across genetic distances

In the population overall, there is a 5.6-fold increased risk of DCM conferred by carrying a TTNtv when adjusting for age, sex, and the first five genetic PCs [**Supplemental Table 2**]. To determine the effect of TTNtvs on DCM risk at different GDs from the European centroid, logistic regression analysis was performed among PMBB individuals binned by GD decile adjusting for age and sex within each bin.

Among those individuals closest to the 1000G European centroid by GD, hiPSI TTNtvs were associated with DCM (Decile 1 OR = 5.82, 95% CI 2.42 to 14.01, *P*<0.001) [**Figure 2A**]; there was no significant effect decomposition as GD decile increased (Decile 10 OR = 9.39, 95% CI 3.82 to 23.13, *P*<0.001). Meta-analysis of binned results demonstrated an overall OR = 5.42 (95% CI 4.01 to 7.32, *P*<0.001); there was no evidence of significant heterogeneity (I^2^=0%, χ^2^=4.59, *P*=0.87). This was supported by a second analysis among all participants in which either GD from the 1000G European centroid alone nor the interaction between GD and hiPSI TTNtvs had a statistically significant effect on the risk of DCM (**Supplemental Table 2**). Taken together these results suggest that GD does not alter DCM risk. Similarly, there was no evident trend of greater or lesser effect on minimum LVEF as GD increased (**Figure 2B**), though there was a trend towards increased heterogeneity (I^2^=42%, χ^2^=15.48, *P*=0.08). We conclude that hiPSI TTNtvs associate with increased DCM risk and reduced minimum LVEF across diverse populations.

**Figure 2:**
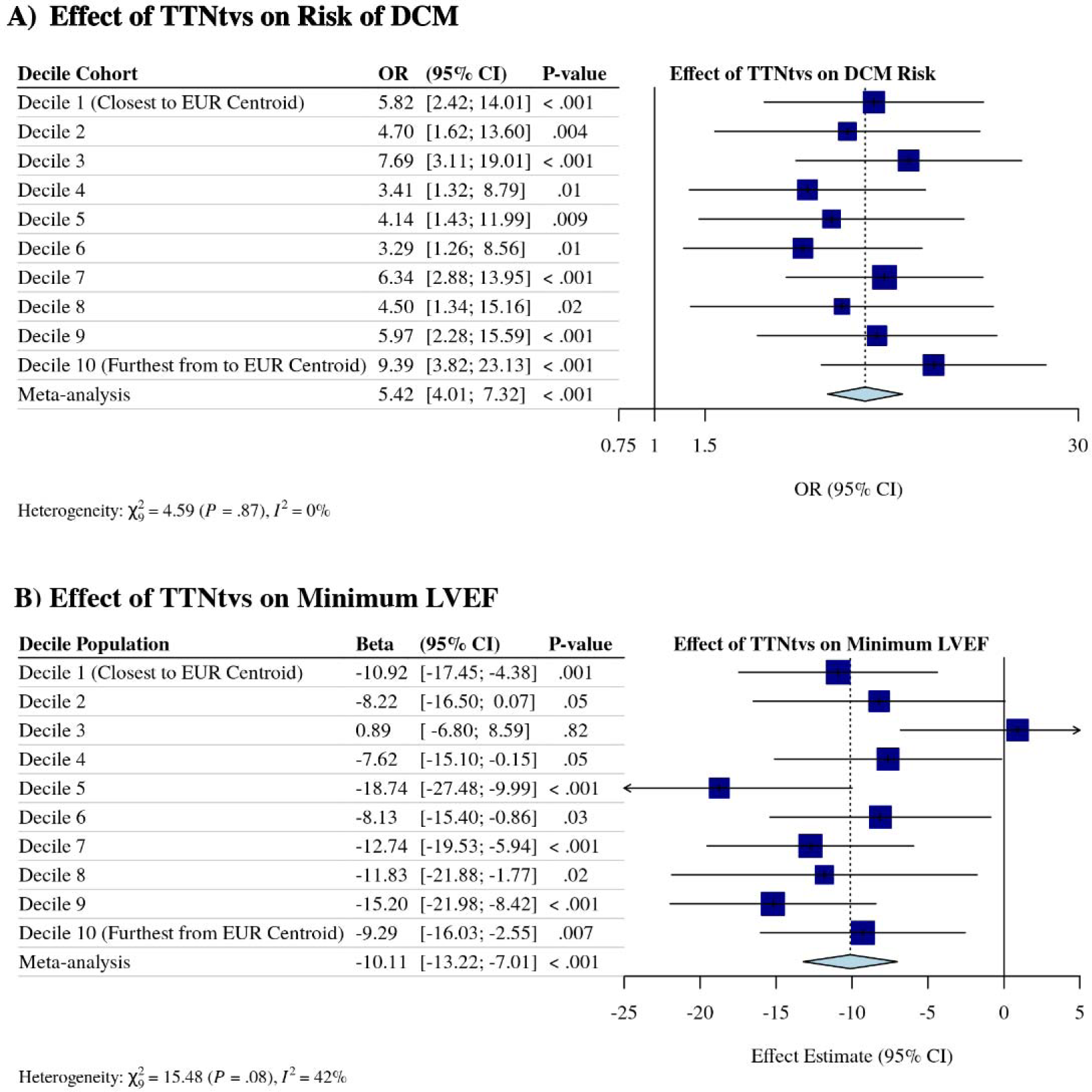
Effect of *TTN* truncating variants on risk of dilated cardiomyopathy and on minimum left ventricular ejection fraction, stratified by genetic distance from the 1000 Genomes Project European centroid. (**A**) Logistic regression analysis of the association between hiPSI TTNtvs on risk of DCM stratified by genetic distance (GD) from the 1000 Genomes Project (1000G) European centroid. (**B**) Linear regression analysis of the association between hiPSI TTNtvs and minimum left ventricular ejection fraction stratified by GD from the 1000G European centroid. OR = odds ratio; CI = confidence interval; EUR = European; LVEF = left ventricular ejection fraction.

### Comparison to categorical assignment of genetic diversity

We previously demonstrated a significant effect on DCM risk conferred by carrying TTNtvs among individuals genetically similar to the 1000G EUR reference population, however could not detect an effect among individuals genetically similar to the 1000G AFR reference population.^7^ In this updated study, encompassing a substantially larger number of participants, we evaluated the effect of TTNtvs among individuals artificially separated into groups to compare both to our previous published results and to the results above. Carrying a hiPSI TTNtv was associated with DCM among individuals genetically similar to the 1000G EUR reference population, whether including (**Supplemental Figure 2**) or excluding individuals with ischemic cardiomyopathy [ICM] (odds ratio [OR] 7.55, 95% CI 4.99 to 11.42, *P*<0.001) [**Figure 3A**]. Similar, but attenuated, results were observed among individuals genetically similar to the 1000G AFR reference population (OR 3.50, 95% CI 1.48 to 8.24, *P*=0.004).

**Figure 3:**
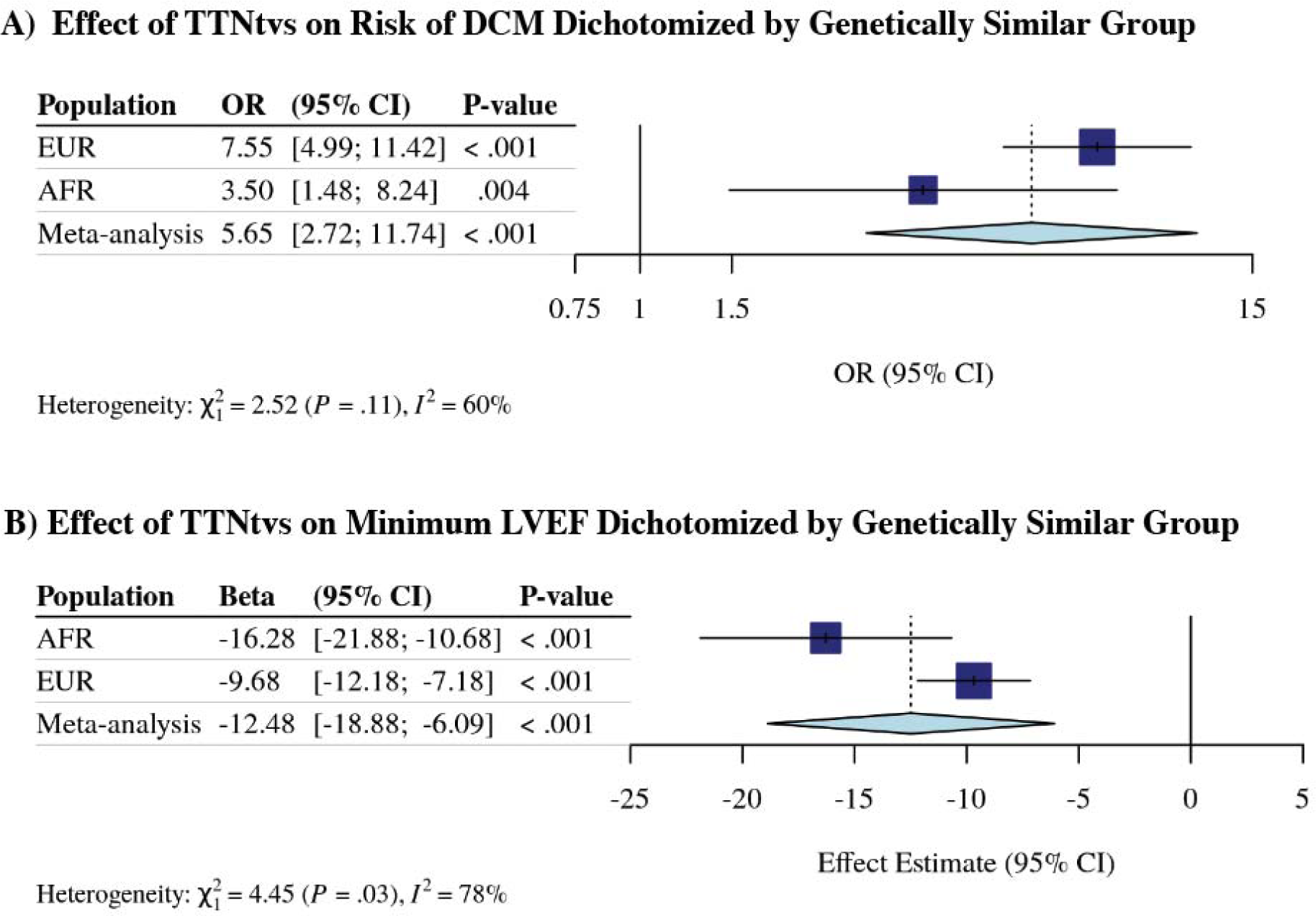
Effect of high percentage spliced in *TTN* truncating variant on risk of dilated cardiomyopathy diagnosis and minimum left ventricular ejection fraction reduction stratified by genetically similar group excluding those with ischemic cardiomyopathy in the Penn Medicine Biobank. (**A**) Logistic regression analysis of the association between hiPSI TTNtvs and DCM diagnosis, and (**B**) linear regression analysis of the association between hiPSI TTNtvs and minimum left ventricular ejection fraction among individuals genetically similar to the 1000 Genomes Project European and African reference population, and meta-analyzed. OR = odds ratio; CI = confidence interval; EUR = individuals genetically similar to the European reference population; AFR = individuals genetically similar to the African reference population.

Linear regression analysis demonstrated a consistent effect on minimum LVEF conferred by carrying a TTNtv without attenuation in the AFR population group [**Figure 3B**]. We conclude that when individuals are artificially grouped by genetic similarity to a reference population, hiPSI TTNtvs continue to associate with a diagnosis of DCM and negatively associate with decreased minimum LVEF across diverse groups.

### Effect of titin truncating variants on risk of atrial fibrillation

To test the effect of TTNtvs on risk of Afib, another cardiovascular diagnosis previously shown to associate with TTNtvs, we used logistic regression analysis by GD and by genetically similar group. Among those individuals closest to the 1000G European centroid by GD, hiPSI TTNtvs were associated with Afib (Decile 1 OR = 2.68, 95% CI 1.40 to 5.16, *P*=0.003) [**Supplemental Figure 3A**]. While there was increased variability across deciles (deciles 2,4, and 5 had statistically insignificant associations between hiPSI TTNtvs and Afib, I^2^=40%, χ^2^=14.96, *P*=0.09), there was no significant trend in effect as GD from the 1000G European centroid increased (Decile 10 OR = 4.36, 95% CI 2.21 to 8.61, *P*<0.001). When PMBB individuals were grouped by genetic similarity to a reference population, the effect estimate among individuals genetically similar to the 1000G EUR reference population (OR = 2.11, 95% CI 1.62 to 2.77, *P*<0.001) was similar to the effect estimate among individuals genetically similar to the 1000G AFR reference population (OR = 1.98, 95% CI 1.02 to 3.85, *P*=0.04) [**Supplemental Figure 3B**]. We conclude that TTNtvs associate with increased risk of Afib across diverse populations.

## Discussion

Using continuous genetic distance as an alternative method to investigate the effect of genetic similarity on rare variant cardiovascular disease risk, we demonstrated that hiPSI TTNtvs associated with increased risk of DCM and reduced minimum LVEF across diverse populations. These results were supported when individuals were artificially dichotomized by genetic similarity to reference populations, although the effect estimate among individuals genetically similar to the AFR reference population may be attenuated. We also demonstrated a consistent though less robust effect of hiPSI TTNtvs on risk of Afib across diverse populations.

Although genetic epidemiology studies have historically relied on arbitrary cutoffs to differentiate and label populations of similar ancestry, genetic ancestry is more appropriately considered as a continuum due to the degree of admixture even among similar populations.^22–24^ The question of TTNtv effect among individuals genetically similar to the 1000G AFR reference population provided an opportunity to reassess the role ancestry plays in how we approach individual risk in cardiovascular disease. The homogeneity of effect of TTNtvs across deciles of GD from the 1000G European centroid removes barriers constructed by presenting results separated by artificially constructed population subgroups, such as within group variation and small sample size. Future evaluation of monogenic risk of cardiovascular disease may benefit from utilizing a similar approach, which may be more broadly applicable than analyses using genetically similar group dichotomization alone.

These results refine our previous findings that hiPSI TTNtvs are associated with DCM among individuals genetically similar to EUR reference populations in PMBB.

Previously, we reported an OR of 18.7 (95% CI 9.1 to 39.4) for DCM among EUR individuals with hiPSI TTNtvs.^7^ The range of effect estimates in our analysis of risk by GD from the 1000G European centroid was OR 3.3 (95% CI 1.3 to 8.6) to OR 9.4 (95% CI 3.8 to 23.1), and the effect estimate for individuals genetically similar to EUR reference panels was contained within that range (OR 7.6, 95% CI 5.0 to 11.4, *P*<0.001). The attenuation of effect we observed between our initial publication and the present study may reflect a winner’s curse in our initial study, where the initially-reported effect size tends to be over-estimated in genetic association studies.^25^ However, the makeup of PMBB has also changed substantially in the past five years such that it is less enriched for cardiovascular disease (6.2% prevalence of DCM previously compared to 2.5% now) due to increasingly broad regional enrollment from centers of primary care which may have had a more meaningful impact on our current results.

Our findings also clarify the effect of hiPSI TTNtvs in individuals genetically similar to the AFR reference population. We previously identified no statistically significant effect among PMBB participants genetically similar to the AFR reference population.^7^ This corroborated other evidence suggesting the possibility of differing effects across diverse populations.^3^ Notably, our previous PMBB cohort included only 2,123 participants genetically similar to the AFR reference population, 20 of whom carried a hiPSI TTNtv, representing a power of 55% to identify an OR of 3.3, suggesting we were previously underpowered to identify an effect similar to what we presently report. We now identify a significant effect of TTNtvs (OR = 3.5, 95% CI 1.5 to 8.2, *P*=0.004) in this population. Moreover, when we consider the extent of admixture that exists in this population, the dispersion of PMBB participants genetically similar to the

1000G AFR reference population across a range of GD from the 1000G European centroid, and the similar effect estimates across those distances, these findings are convincing that hiPSI TTNtvs confer increased DCM risk among individuals within these GD deciles corresponding to those previously labeled genetically similar to the 1000G AFR reference population.

## Limitations

This work has some limitations. Reliance on electronic health records to establish a diagnosis of DCM may have misclassified a subset of individuals. Similarly, TTEs were not interpreted by a uniform reader. Our cohort is also largely composed of EUR and AFR individuals, limiting generalizability of the GD continuum concept to other genetically similar groups.

## Conclusion

In summary, our updated analysis demonstrates a strong association between hiPSI TTNtvs and DCM across a range of genetic backgrounds, including subjects genetically similar to African reference population. These data suggest that recommendations for genetic testing and counseling should not differ between diverse populations.

## Data Availability

All data produced in the present study are available upon reasonable request to the authors.

## Acknowledgements, Conflicts of Interest and Disclosures

We acknowledge the Penn Medicine BioBank (PMBB) for providing data and thank the patient-participants of Penn Medicine who consented to participate in this research program. We would also like to thank the Penn Medicine BioBank team and Regeneron Genetics Center for providing genetic variant data for analysis. The PMBB is approved under IRB protocol# 813913 and supported by Perelman School of Medicine at University of Pennsylvania, a gift from the Smilow family, and the National Center for Advancing Translational Sciences of the National Institutes of Health under CTSA award number UL1TR001878. M.G.L. receives research support to his institution from MyOme outside of this work. S.M.D receives research support to his institution from RenalytixAI and in kind support from Novo Nordisk, both outside of this work.

## Sources of Funding

J.D. is supported by the American Heart Association (23POST1011251). M.G.L. received support from the Institute for Translational Medicine and Therapeutics of the Perelman School of Medicine at the University of Pennsylvania, the NIH/NHLBI National Research Service Award postdoctoral fellowship (T32HL007843), the Measey Foundation, and the Doris Duke Foundation (2023-0224). Z.A. is supported by the NIH/NHLBI (R01-HL152446) and the Department of Defense (W81XWH18-1-0503).

## Notes

### Competing Interest Statement

The authors have declared no competing interest.

### Funding Statement

Funding for this study included the following: JD was funded by American Heart Association (23POST1011251). M.G.L. received support from the Institute for Translational Medicine and Therapeutics of the Perelman School of Medicine at the University of Pennsylvania, the NIH/NHLBI National Research Service Award postdoctoral fellowship (T32HL007843), the Measey Foundation, and the Doris Duke Foundation (2023-0224). Z.A. is supported by the NIH/NHLBI (R01-HL152446) and the Department of Defense (W81XWH18-1-0503).

### Author Declarations

This work was approved by the Univeersity of Pennsylvania Institutional Review Board, protocol# 813913.

## References

1. Freiburg A, Trombitas K, Hell W, Cazorla O, Fougerousse F, Centner T, Kolmerer B, Witt C, Beckmann JS, Gregorio CC, et al. Series of exon-skipping events in the elastic spring region of titin as the structural basis for myofibrillar elastic diversity. Circ Res. 2000;86:1114–1121. doi: 10.1161/01.res.86.11.1114

2. Herman DS, Lam L, Taylor MR, Wang L, Teekakirikul P, Christodoulou D, Conner L, DePalma SR, McDonough B, Sparks E, et al. Truncations of titin causing dilated cardiomyopathy. N Engl J Med. 2012;366:619–628. doi: 10.1056/NEJMoa1110186

3. Roberts AM, Ware JS, Herman DS, Schafer S, Baksi J, Bick AG, Buchan RJ, Walsh R, John S, Wilkinson S, et al. Integrated allelic, transcriptional, and phenomic dissection of the cardiac effects of titin truncations in health and disease. Sci Transl Med. 2015;7:270ra276. doi: 10.1126/scitranslmed.3010134

4. Akinrinade O, Alastalo TP, Koskenvuo JW. Relevance of truncating titin mutations in dilated cardiomyopathy. Clin Genet. 2016;90:49–54. doi: 10.1111/cge.12741

5. Haas J, Frese KS, Peil B, Kloos W, Keller A, Nietsch R, Feng Z, Muller S, Kayvanpour E, Vogel B, et al. Atlas of the clinical genetics of human dilated cardiomyopathy. Eur Heart J. 2015;36:1123–1135a. doi: 10.1093/eurheartj/ehu301

6. Akinrinade O, Ollila L, Vattulainen S, Tallila J, Gentile M, Salmenpera P, Koillinen H, Kaartinen M, Nieminen MS, Myllykangas S, et al. Genetics and genotype- phenotype correlations in Finnish patients with dilated cardiomyopathy. Eur Heart J. 2015;36:2327–2337. doi: 10.1093/eurheartj/ehv253

7. Haggerty CM, Damrauer SM, Levin MG, Birtwell D, Carey DJ, Golden AM, Hartzel DN, Hu Y, Judy R, Kelly MA, et al. Genomics-First Evaluation of Heart Disease Associated With Titin-Truncating Variants. Circulation. 2019;140:42–54. doi: 10.1161/CIRCULATIONAHA.119.039573

8. McAfee Q, Chen CY, Yang Y, Caporizzo MA, Morley M, Babu A, Jeong S, Brandimarto J, Bedi KC, Jr., Flam E, et al. Truncated titin proteins in dilated cardiomyopathy. Sci Transl Med. 2021;13:eabd7287. doi: 10.1126/scitranslmed.abd7287

9. McAfee Q, Caporizzo MA, Uchida K, Bedi KC, Jr., Margulies KB, Arany Z, Prosser BL. Truncated titin protein in dilated cardiomyopathy incorporates into the sarcomere and transmits force. J Clin Invest. 2023. doi: 10.1172/JCI170196

10. Jordan E, Kinnamon DD, Haas GJ, Hofmeyer M, Kransdorf E, Ewald GA, Morris AA, Owens A, Lowes B, Stoller D, et al. Genetic Architecture of Dilated Cardiomyopathy in Individuals of African and European Ancestry. JAMA. 2023;330:432–441. doi: 10.1001/jama.2023.11970

11. Astore C, Sharma S, Nagpal S, Consortium NIG, Cutler DJ, Rioux JD, Cho JH, McGovern DPB, Brant SR, Kugathasan S, et al. The role of admixture in the rare variant contribution to inflammatory bowel disease. Genome Med. 2023;15:97. doi: 10.1186/s13073-023-01244-w

12. Lewis ACF, Molina SJ, Appelbaum PS, Dauda B, Di Rienzo A, Fuentes A, Fullerton SM, Garrison NA, Ghosh N, Hammonds EM, et al. Getting genetic ancestry right for science and society. Science. 2022;376:250–252. doi: 10.1126/science.abm7530

13. Busby GB, Kulm S, Bolli A, Kintzle J, Domenico PD, Botta G. Ancestry-specific polygenic risk scores are risk enhancers for clinical cardiovascular disease assessments. Nat Commun. 2023;14:7105. doi: 10.1038/s41467-023-42897-w

14. Ding Y, Hou K, Xu Z, Pimplaskar A, Petter E, Boulier K, Prive F, Vilhjalmsson BJ, Olde Loohuis LM, Pasaniuc B. Polygenic scoring accuracy varies across the genetic ancestry continuum. Nature. 2023;618:774–781. doi: 10.1038/s41586-023-06079-4

15. Ahlberg G, Refsgaard L, Lundegaard PR, Andreasen L, Ranthe MF, Linscheid N, Nielsen JB, Melbye M, Haunso S, Sajadieh A, et al. Rare truncating variants in the sarcomeric protein titin associate with familial and early-onset atrial fibrillation. Nat Commun. 2018;9:4316. doi: 10.1038/s41467-018-06618-y

16. Corden B, Jarman J, Whiffin N, Tayal U, Buchan R, Sehmi J, Harper A, Midwinter W, Lascelles K, Markides V, et al. Association of Titin-Truncating Genetic Variants With Life-threatening Cardiac Arrhythmias in Patients With Dilated Cardiomyopathy and Implanted Defibrillators. JAMA Netw Open. 2019;2:e196520. doi: 10.1001/jamanetworkopen.2019.6520

17. Akhtar MM, Lorenzini M, Cicerchia M, Ochoa JP, Hey TM, Sabater Molina M, Restrepo-Cordoba MA, Dal Ferro M, Stolfo D, Johnson R, et al. Clinical Phenotypes and Prognosis of Dilated Cardiomyopathy Caused by Truncating Variants in the TTN Gene. Circ Heart Fail. 2020;13:e006832. doi: 10.1161/CIRCHEARTFAILURE.119.006832

18. Dewey FE, Gusarova V, O’Dushlaine C, Gottesman O, Trejos J, Hunt C, Van Hout CV, Habegger L, Buckler D, Lai KM, et al. Inactivating Variants in ANGPTL4 and Risk of Coronary Artery Disease. N Engl J Med. 2016;374:1123–1133. doi: 10.1056/NEJMoa1510926

19. Verma A, Damrauer SM, Naseer N, Weaver J, Kripke CM, Guare L, Sirugo G, Kember RL, Drivas TG, Dudek SM, et al. The Penn Medicine BioBank: Towards a Genomics-Enabled Learning Healthcare System to Accelerate Precision Medicine in a Diverse Population. J Pers Med. 2022;12. doi: 10.3390/jpm12121974

20. Wang K, Li M, Hakonarson H. ANNOVAR: functional annotation of genetic variants from high-throughput sequencing data. Nucleic Acids Res. 2010;38:e164. doi: 10.1093/nar/gkq603

21. Fairley S, Lowy-Gallego E, Perry E, Flicek P. The International Genome Sample Resource (IGSR) collection of open human genomic variation resources. Nucleic Acids Res. 2020;48:D941–D947. doi: 10.1093/nar/gkz836

22. Mathias RA, Taub MA, Gignoux CR, Fu W, Musharoff S, O’Connor TD, Vergara C, Torgerson DG, Pino-Yanes M, Shringarpure SS, et al. A continuum of admixture in the Western Hemisphere revealed by the African Diaspora genome. Nat Commun. 2016;7:12522. doi: 10.1038/ncomms12522

23. Tishkoff SA, Reed FA, Friedlaender FR, Ehret C, Ranciaro A, Froment A, Hirbo JB, Awomoyi AA, Bodo JM, Doumbo O, et al. The genetic structure and history of Africans and African Americans. Science. 2009;324:1035–1044. doi: 10.1126/science.1172257

24. Gouveia MH, Borda V, Leal TP, Moreira RG, Bergen AW, Kehdy FSG, Alvim I, Aquino MM, Araujo GS, Araujo NM, et al. Origins, Admixture Dynamics, and Homogenization of the African Gene Pool in the Americas. Mol Biol Evol. 2020;37:1647–1656. doi: 10.1093/molbev/msaa033

25. Lohmueller KE, Pearce CL, Pike M, Lander ES, Hirschhorn JN. Meta-analysis of genetic association studies supports a contribution of common variants to susceptibility to common disease. Nat Genet. 2003;33:177–182. doi: 10.1038/ng1071

