## Supplemental Data File for "Titin-Truncating variants Predispose to Dilated Cardiomyopathy in Diverse Populations"

### Tables and Figure Legends

**Supplemental Table 1: Number of Individuals Genetically Similar to a 1000 Genomes Project Reference Population in Each Decile of Genetic Distance from the European Centroid.**

| GD Decile from European Centroid | EUR | AFR | AMR | SAS | EAS |
|---|---|---|---|---|---|
| 1 | 4226 | 42 | 1 | 0 | 0 |
| 2 | 4134 | 133 | 2 | 0 | 0 |
| 3 | 3814 | 447 | 8 | 0 | 0 |
| 4 | 3246 | 1013 | 10 | 0 | 0 |
| 5 | 2579 | 1679 | 10 | 1 | 0 |
| 6 | 2172 | 2085 | 10 | 2 | 0 |
| 7 | 2110 | 2134 | 21 | 4 | 0 |
| 8 | 2476 | 1734 | 49 | 9 | 0 |
| 9 | 2948 | 1205 | 95 | 20 | 0 |
| 10 | 2251 | 664 | 360 | 523 | 470 |

GD = Genetic Distance; EUR = 1000 Genomes Project European Reference Population; AFR = 1000 Genomes Project African Reference Population; AMR = 1000 Genomes Project Ad Mixed American Reference Population; SAS = 1000 Genomes Project South Asian Reference Population; EAS = 1000 Genomes Project East Asian Reference Population.

**Supplemental Table 2: Effect of hiPSI *TTN* truncating variants, genetic distance from 1000 Genomes Project European centroid, and the interaction between genetic distance and hiPSI *TTN* truncating variants on risk of dilated cardiomyopathy.**

| **Regression Variable** | **Odds Ratio of DCM** | **95% Confidence Interval** | ***P*-value** |
|---|---|---|---|
| hiPSI TTNtv | 5.63 | 4.16 to 7.61 | <0.001 |
| GD | $7.84 \times 10^{-8}$ | $1.63 \times 10^{-16}$ to 23.30 | 0.10 |
| hiPSI TTNtv*GD | $1.84 \times 10^{24}$ | $4.09 \times 10^{-9}$ to $8.27 \times 10^{56}$ | 0.15 |

hiPSI = high percentage spliced in; DCM = Dilated cardiomyopathy; GD = Genetic Distance from the 1000 Genomes Project European Centroid.

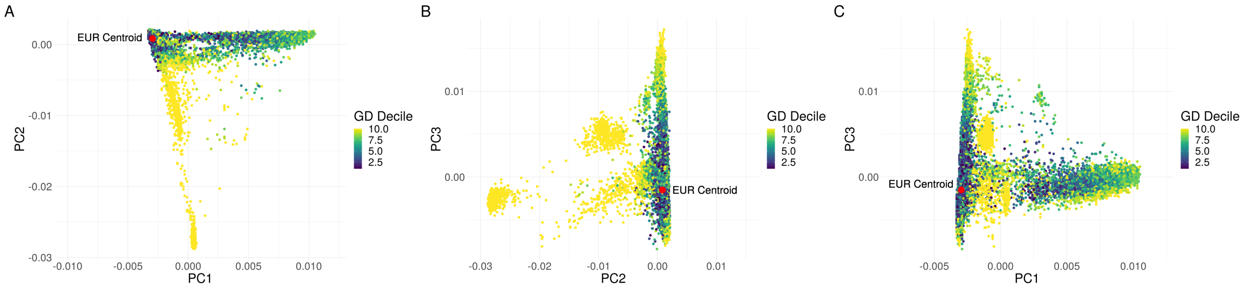


**Supplemental Figure 1: Principal component based clusters of individuals in the Penn Medicine Biobank compared to genetic distance from the 1000 Genomes Project European centroid.** Discrete labelling of the position of each individual within PMBB colored by decile of genetic distance from the 1000 Genomes Project (1000G) European centroid in a plot of (**A**) principal component (PC) 1 versus PC2, (**B**) PC2 versus PC3, and (**C**) PC1 versus PC3 with 1000G European centroid (red dot) identified in each plot demonstrating the overlap between individuals genetically close or distant from the 1000G European centroid based on PC space.

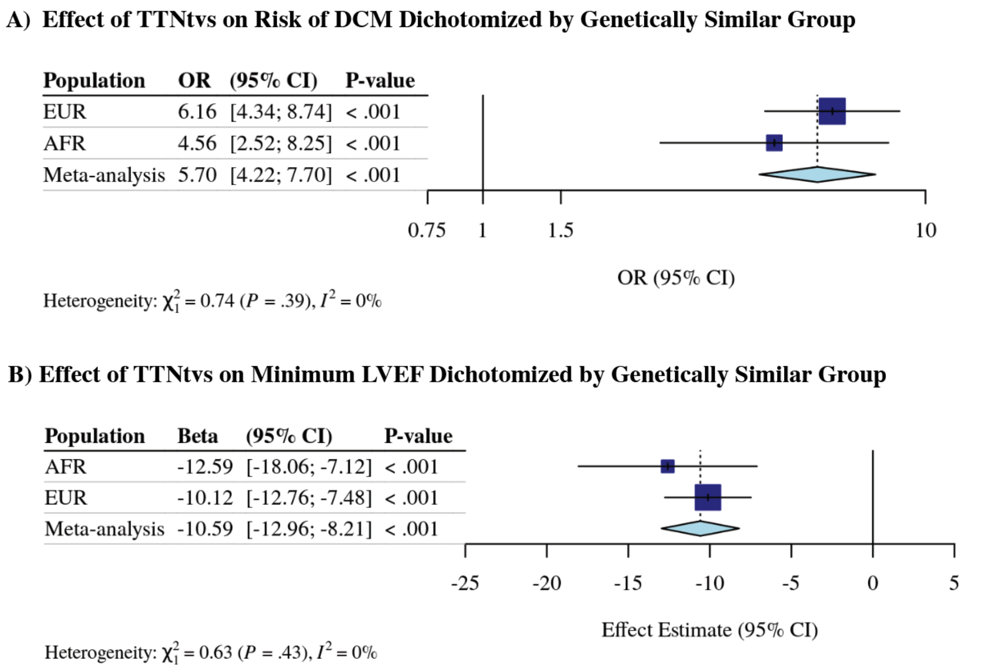


**Supplemental Figure 2: Effect of high percentage spliced in titin truncating variant on risk of dilated cardiomyopathy diagnosis and minimum left ventricular ejection fraction reduction stratified by genetically similar group without excluding those with ischemic cardiomyopathy in the Penn Medicine Biobank**. (**A**) Logistic regression analysis of the association between hiPSI TTNtvs and DCM diagnosis, and (**B**) linear regression analysis of the association between hiPSI TTNtvs and minimum left ventricular ejection fraction among individuals genetically similar to the 1000 Genomes Project European and African reference population, and meta-analyzed. OR = odds ratio; CI = confidence interval; EUR = individuals genetically similar to the European reference population; AFR = individuals genetically similar to the African reference population.

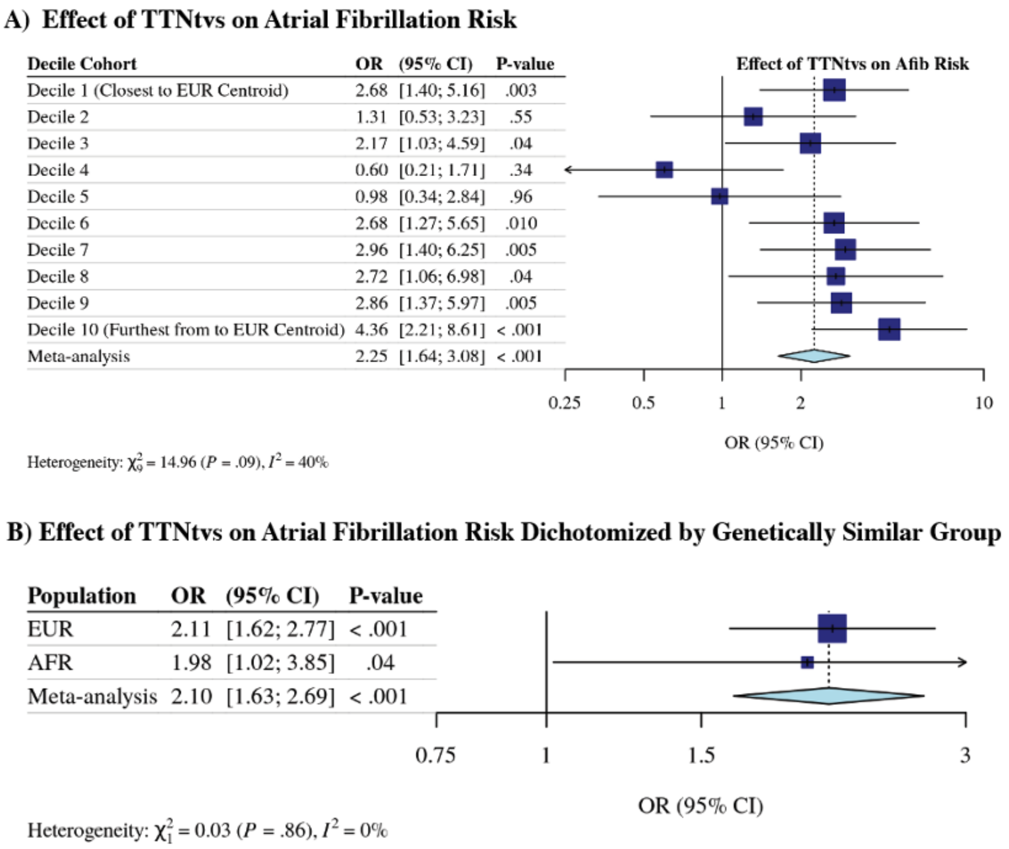


**Supplemental Figure 3: Effect of high percentage spliced in titin truncating variant on risk of atrial fibrillation diagnosis in the Penn Medicine Biobank**. (**A**) Logistic regression analysis of the association between hiPSI TTNtvs and Afib by deciles of genetic distance from the 1000 Genomes Project European centroid; and (**B**) logistic regression analysis of the association between hiPSI TTNtvs and Afib by genetically similar group. OR = odds ratio; CI = confidence interval; EUR = individuals genetically similar to the European reference population; AFR = individuals genetically similar to the African reference population.
